# Presentation of long COVID and associated risk factors in a mobile health study

**DOI:** 10.1101/2022.09.27.22280404

**Authors:** Callum Stewart, Yatharth Ranjan, Pauline Conde, Shaoxiong Sun, Zulqarnain Rashid, Heet Sankesara, Nicholas Cummins, Petroula Laiou, Xi Bai, Richard J B Dobson, Amos A Folarin

**Affiliations:** Department of Health Informatics and Biostatistics, Institute of Psychiatry, Psychology and Neuroscience, Kings College London, UK; Institute of Health Informatics, University College London, UK

## Abstract

**Backgroun:** The Covid Collab study was a citizen science mobile health research project set up in June 2020 to monitor COVID-19 symptoms and mental health through questionnaire self-reports and passive wearable device data.

**Methods:** Using mobile health data, we consider whether a participant is suffering from long COVID in two ways. Firstly, by whether the participant has a persistent change in a physiological signal commencing at a diagnosis of COVID-19 that last for at least twelve weeks. Secondly, by whether a participant has self-reported persistent symptoms for at least twelve weeks. We assess sociodemographic and wearable-based risk factors for the development of long COVID according to the above two categorisations.

**Findings:** Persistent changes to physiological signals measured by commercial fitness wearables, including heart rate, sleep, and activity, are visible following a COVID-19 infection and may help differentiate people who develop long COVID. Anxiety and depression are significantly and persistently affected at a group level following a COVID-19 infection. We found the level of activity undertaken in the year prior to illness was protective against long COVID and that symptoms of depression before and during the acute illness may be a risk factor.

**Interpretation:** Mobile health and wearable devices may prove to be a useful resource for tracking recovery and presence of long-term sequelae to COVID-19. Mental wellbeing is significantly negatively effected on average for an extended period of time following a COVID-19 infection.

## Introduction

There have been over 500 million confirmed severe acute respiratory syndrome coronavirus 2 (SARS-CoV-2) infections as of April 2022 [1]. Despite the development and successful rollout of vaccinations and treatments, COVID-19 remains a danger both in terms of the acute illness, chance of death, long-term illness following infection, and the possibility of problematic variants developing.

Persistent symptoms following a SARS-CoV-2 infection, often termed *long COVID* or *Post Acute COVID-19 syndrome*, are thought to affect a significant number of patients. The presence of these long-term effects was largely illuminated early in the pandemic through subjective accounts from patients of the disease [2]. While there has been a flurry of research starting to address the prevalence, clinical features, and risk factors [3] of long COVID, our understanding of the condition remains sparse.

Attempts have been made to categorise long COVID (LCOVID) on the basis of symptomatology, time periods, and aetiology, but it remains a loosely defined syndrome with multiple associated terminologies [4]. In the United Kingdom, the National Institute for Health and Care Excellence (NICE) suggest the use of *Acute COVID-19* for symptoms up to four weeks, *Ongoing symptomatic COVID-19* for symptoms from four to twelve weeks, and *Post-COVID-19 syndrome* for symptoms continuing past twelve weeks[5]. Symptoms have been reported across a wide range of organs and body systems, including cardiorespiratory, neurological, psychological, musculatory, gastrointestinal, and systemic[6, 7]. Common symptoms include but are not limited to fatigue, dyspnea, anxiety, sleep disorders, pain, dizziness, and anosmia.

Prevalence estimates for LCOVID vary, partially with respect to study cohorts, terminology, and study design. Many studies recruit from hospitalised populations and therefore select for severe cases of COVID-19. A recent large-scale community study on self-reported symptoms estimated [3.1-5.8]% of participants experienced at least one persistent symptom for over 12 weeks following a COVID infection [3].

The pandemic has been a focal point for the greater emergence of digital health technologies in research and healthcare. Within COVID-19 research, there are multiple large studies using digital health and ‘big data’ approaches to better understand trajectories of, diagnose, and estimate the prevalence of COVID-19 and its long-term sequelae. Mobile health (mHealth) data modalities can offer an insight into LCOVID complementary to existing studies. As a scalable and continuous data collection method, passive mobile health sensing provides an objective measure of health. Additionally, long periods of historical data is often available from wearable fitness devices. The availability of wearable data outside of medical care pathways grants an avenue to observe people who may otherwise be missed. For example, significant burdens to health and function in people with flu who do not seek medical care because of the sub-clinical nature of their illness have been demonstrated by making use of commercial wearable sensor data [8].

Covid Collab is an observational mHealth study which began in June 2020. Participants enrolled through the study app, Mass Science, and were prompted to complete regular surveys on COVID symptoms experienced, vaccination and diagnosis status, mood, and mental well-being. Participants were able to share existing and prospective wearable data through their Fitbit and Garmin accounts[9]. We collect wearable data covering a period prior to the pandemic, giving a historic baseline against which to compare. Additionally, we regularly prompted for the completion of self-reported questionnaires on current symptoms and mental health throughout the study, providing a contemporaneous account of mental wellbeing and COVID-19-associated symptoms before, throughout, and after any COVID-19 infection.

We queried Pubmed from inception up until 01 Jul 2022 for studies on LCOVID that use wearable or mHealth technologies using the string ‘((COVID* OR SARS-COV-2) AND (long OR persistent OR hauler OR post OR sequelae)) AND (mHealth OR wearable OR telemedicine OR app)’. Of the 2144 results, the vast majority of returned studies concerned the role of telemedicine in delivering care of other conditions since the start of the COVID-19 pandemic. A number of studies relate to the monitoring, detection, or diagnosis of acute COVID-19. Others use digital technology or telemedicine in the treatment or to assess the impact of rehabilitation courses. Eight studies investigate LCOVID through remote digital technologies. Of those, six used self-reported questionnaire data collected through televisits or apps to characterise symptomatology and trajectories of LCOVID, including a large-scale community study from the ZOE COVID Symptom app. Three studies use passively collected data from commercial or experimental wearable sensors to describe how heart rate, sleep, and activity change in a COVID-19 positive population following infection. Two of those show a pattern of bradycardia and tachycardia in resting heart rate, with a persistent change in some cases lasting over four months.

Covid Collab provides a unique viewpoint for quantifying the features and risk factors of LCOVID. This study incorporates survey data alongside pervasive wearable sensor data. Participant self-reported questionnaires included regular mental health measures as well as physical COVID-19 related symptoms. The study population was recruited remotely and openly throughout the pandemic. It therefore includes non-hospitalised participants and those with a mild response to acute COVID-19 infection, with data often collected prior to infection which does not rely on participant recall and the inclusion of historic wearable data from prior to enrolment. Software and data collection infrastructure have been open sourced to facilitate the reuse of this system for future digital epidemiology research or monitoring programmes.

The aims of this paper are to: (1) Quantify the prevalence and severity of long-term symptoms across collected mHealth metrics including heart rate, sleep, physical activity, and self-reported symptoms and mood. (2) Identify risk factors for the severity and duration of persistent symptoms. Accordingly LCOVID (LCOVID) is considered here as persistent changes or symptoms at and beyond the twelfth week post-diagnosis.

## Methods

### Study design and participants

The study is a longitudinal self-enrolled community study administered through a smartphone app. As of August 2022, 17,667 participants had enrolled. Participation was open to any adult greater than or equal to 18 years of age. However, enrolment was skewed towards those based in the United Kingdom (UK) and of female gender (N=12137, 68.7%) [Table 1]. Recruitment was carried out via the study app, media publications, and promotion within the UK version of the Fitbit app between August 2020 and May 2021.

**Table 1:** Sociodemographic statistics in Covid Collab

| Demographic category | Cases | Controls |
| --- | --- | --- |
| **Initial questionnaire** |  |  |
| Age [min-max] | 44.78±13.18 [15.0-87.0] | 48.30±13.28 [15.00-88.00] |
| Sex = f | 1247 (76.1%) | 2682 (74.5%) |
| Height [min-max] (cm) | 168.24±9.83 [118.0-220.0] | 168.59±9.79 [100.00-212.00] |
| Weight [min-max] (kg) | 78.45±18.43 [26.50-146.20] | 77.14±17.57 [31.70-148.30] |
| **Smoking** |  |  |
| Non-smoker | 917 (56.71%) | 2118 (58.87%) |
| Ex-smoker | 461 (28.51%) | 1030 (28.63%) |
| Current smoker | 239 (14.78%) | 450 (12.51%) |
| **Comorbidities (Extended questionnaire, cases n=759, control n=2229)** |  |  |
| Asthma | 173 (22.79%) | 395 (17.72%) |
| Hypertension | 83 (10.94%) | 256 (11.48%) |
| Diabetes | 31 (4.08%) | 88 (3.95%) |
| Depression | 173 (22.79%) | 532 (23.87%) |
| Anxiety | 136 (17.92%) | 356 (15.97%) |
| **Employment (Extended questionnaire)** |  |  |
| Full time | 411 (54.15%) | 1079 (48.41%) |
| Part time | 125 (16.47%) | 321 (14.40%) |
| Retired | 97 (12.78%) | 448 (20.10%) |
| Student | 22 (2.90%) | 72 (3.23%) |
| Unemployed | 19 (2.50%) | 57 (2.56%) |
| **Marital and family status (Extended questionnaire)** |  |  |
| In a relationship | 592 (78.72%) | 1697 (77.17%) |
| Single or separated | 156 (20.74%) | 498 (22.65%) |
| Unknown | 4 (0.53%) | 4 (0.18%) |
| With children | 553 (73.54%) | 1413 (64.26%) |
| **Living Situation (Extended questionnaire)** |  |  |
| Alone | 61 (8.04%) | 306 (13.73%) |
| With partner | 454 (59.82%) | 1428 (64.06%) |
| With family | 255 (33.60%) | 523 (23.46%) |
| With children | 204 (26.88%) | 411 (18.44%) |
| With adult children | 122 (16.07%) | 240 (10.77%) |
| Houseshare | 20 (2.64%) | 57 (2.56%) |

Sociodemographic data from initial enrolment and an extended questionnaire across the COVID-positive cohort and the control group of COVID-negative participants.

Participants were included in the analysis if they reported a COVID-19 diagnosis (with antibody or PCR tests) before 2022/02/01. In total, 1,743 participants were included. Different numbers of participants were included in different aspects of the analysis because of the differing rates of completion between modalities. A group of age-, sex-, and time-matched controls (N=3,600) were selected from participants with high questionnaire completion rates who had not reported a COVID-19 diagnosis.

A detailed explanation of the study protocol in Covid Collab has previously been published [9]. The development of the pandemic and our changing understanding of the disease and requirements of the study led to some amendments to the protocol, including an extended socio-demographics questionnaire building on the initial registration questionnaire. Additionally, participants were able to donate from different sources of data, but were free to provide as much as they chose. Because of those two reasons, there are differences in data availability between participants. Of the participants included in this study, 759 (43.5%) had completed the extended socio-demographic questionnaire.

### Study metrics

There are two major categories of data in this study: passive and active data (Table 2). Active data refers to questionnaires delivered in-app that require conscious participant engagement. The questionnaires include the PHQ-8 scale of depression[10], the GAD-7 scale of generalised anxiety[11], a visual analog scale for arousal and valence[12], and a COVID-19 symptoms questionnaire. In addition, participants are able to submit COVID-19 diagnosis (antigen, PCR, or symptom determined) and vaccination events. Passive data refers to data collected from instruments that do not require any conscious participant involvement. In this study, the passive metrics consist of heart rate, heart rate variability, sleep, step counts, and activity logs provided through the participant’s sharing of commercial wearable sensor data.

**Table 2:** Active and passive mobile health metrics collected in this study.

| Category | Metric | Frequency | Description |
| --- | --- | --- | --- |
| Questionnaires | PHQ-8 | 14 days | [10] |
|  | GAD-7 | 14 days | [11] |
|  | Arousal-Valence | Ad-hoc / twice-weekly | [12] |
|  | Symptoms relating to COVID-19 | Ad-hoc / twice-weekly |  |
|  | Diagnosis | Ad-hoc | Self report diagnosis by Antigen, PCR, Symptom |
| Fitbit | Heart rate | Daily | Daily resting heart rate provided by Fitbit Web & API.[13] |
|  | Heart rate variability | Daily | Daily root mean square of successive differences (RMSSD) provided by Fitbit Web API[14] |
|  | Sleep duration | Per-sleep | Estimated duration of a sleep[15] |
|  | Sleep efficiency | Per-sleep | Fitbit calculates an 'efficiency' score for each recorded sleep.[15] |
|  | Step count | Daily | [15] |
|  | Activity log | Ad-hoc | [15] |

### Group analysis

A comparison of passive and self-reported metrics between case and control groups was carried out. Group-wide resting heart rate (RHR), heart rate variability (HRV) measured as the Fitbit daily root mean square of successive differences (dailyRMSSD), sleep duration, sleep efficiency, step count, PHQ-8 score, GAD-7 score, and self-rated valence and arousal are compared at acute (<4 weeks), ongoing (4-12 weeks), and post-COVID (>12 weeks) syndrome periods, as defined in NICE guidelines[5], following a self-reported diagnosis of COVID-19 to a time-matched group of control participants. Analysis is carried out in Python. A Brunner-Munzul test was carried out between the two groups for each set of metrics using the statsmodels library[16]. Self-reported symptom counts, severity and durations were visualised to understand the symptomatology of COVID-19 and to explore long lasting symptoms.

### Risk factors for LCOVID

To test risk factors for LCOVID it is necessary to define a candidate group of participants who are likely to have LCOVID on the basis of the data that we have available. We consider two approaches.

Firstly, we consider using the change in resting heart rate over a period of 12 weeks post-COVID-19 infection as a proxy for LCOVID (the RHR-LCOVID cohort), where a greater change compared to a baseline would indicate a more likely case or greater severity of LCOVID. In this categorisation, we do not explicitly group participants but instead use the change in heart rate as a continuous outcome variable. To do so, we need to estimate a baseline predicted heart rate that the participant would be expected to have if they did not have a COVID-19 infection. To estimate an expected resting heart rate, we fit a Bayesian structural time series model on each participant’s resting heart rate up until the date of diagnosis using the CausalImpact library[17]. The model comprises a local-level model, a seasonal model with a period of 28 days, and a regularised regression on a set of 500 participants who were not otherwise involved in the analysis. The change in resting heart rate at 12 weeks is used as the outcome in a linear regression with age, sex, historic activity, historic sleep duration, and the change in RHR between the baseline and acute phase. The historic activity and sleep duration are taken from Fitbit data between one and two years prior to diagnosis. The historic activity is the average duration of time in minutes spent in the Fitbit ‘high activity’ level per day. The sleep duration is the average time spent asleep per day. The ‘historic’ period is taken as the period two years until half a year before diagnosis of COVID-19. The baseline to acute change in RHR is defined as the difference between the average RHR 1-4 weeks prior to a COVID-19 diagnosis and 0-4 weeks after a COVID-19 diagnosis.

Secondly, we consider participants who have self-reported symptoms for an extended period following a self-reported diagnosis of COVID-19. The self-reported symptoms submitted by all participants who reported a positive diagnosis were used to determine length of illness and split the diagnosed cohort into short- and long-COVID groups. If at least one symptom was reported at least once per week for at least twelve weeks, the participant is assigned to the symptom-based LCOVID group (*L*_*symp*_). Participants were otherwise assigned to the symptom-based short COVID group (*S*_*symp*_). Risk factor assessment using logistic regression was performed on the *S*_*symp*_ and *L*_*symp*_ groups based on demographics, baseline passive data, and mental health scores during the acute phase of COVID infection.

## Results

### Group-wide analysis

Passive wearable device metrics and self-reported mental health survey scores were compared between case (COVID-19+) and control groups at three time points (Table 3). The ‘acute’ period was defined as between the date of diagnosis and four weeks post-diagnosis. The ‘ongoing’ period took values between four weeks and eight weeks post-diagnosis. The ‘post-COVID’ period took values between twelve and sixteen weeks post-diagnosis. For each period and metric, a comparison of the case and control distributions of the mean values for each of the constituent participants was carried out. A Brunner-Munzul two group non-parametric test[18] was calculated to compare each distribution. Significance was determined with a p-value cutoff of 0.05 after Benjamini-Hochberg correction[19].

**Table 3:** Group-wide Comparisons

| Metric | Acute (<4w) |  |  | Ongoing (4-12w) |  |  | Post-COVID (>12w) |  |  |
| --- | --- | --- | --- | --- | --- | --- | --- | --- | --- |
|  | Case | Control | p-value | Case | Control | p-value | Case | Control | p-value |
| RHR | 0.69<br>± 3.44 | 0.11<br>± 3.15 | **6.97e-05** | 1.17<br>± 3.35 | 0.07<br>± 3.19 | **1.17e-16** | 0.63<br>± 3.44 | 0.17<br>± 3.28 | **3.62e-04** |
| RMSSD | 13.58<br>± 0.38 | 13.58<br>± 0.39 | 0.79 | 13.67<br>± 0.43 | 13.68<br>± 0.44 | 0.75 | 13.62<br>± 0.39 | 13.62<br>± 0.39 | 0.99 |
| Steps | -1478<br>± 3635 | 39.44<br>± 3572 | **6.74e-37** | -49.2<br>± 3820 | 128<br>± 3630 | 0.42 | 288<br>± 3828 | 464<br>± 3691 | 0.93 |
| Sleep efficiency | -1.34<br>± 7.09 | -0.83<br>± 6.30 | **5.05e-06** | -1.11<br>± 6.75 | -0.81<br>± 6.47 | **0.03** | -1.23<br>± 7.72 | -0.98<br>± 6.58 | **1.29e-03** |
| Sleep duration | 5.98<br>± 63.06 | 4.88<br>± 56.07 | 0.67 | 6.90<br>± 65.34 | 4.53<br>± 59.25 | 0.73 | 2.81<br>± 70.34 | 2.49<br>± 60.78 | 0.90 |
| PHQ-8 | 7.96<br>± 6.00 | 5.22<br>± 5.30 | **4.90e-29** | 6.98<br>± 6.00 | 5.29<br>± 5.35 | **1.13e-07** | 6.21<br>± 5.57 | 5.23<br>± 5.48 | **1.53e-03** |
| GAD-7 | 5.88<br>± 5.36 | 4.49<br>± 5.02 | **1.03e-10** | 5.34<br>± 5.27 | 4.60<br>± 5.07 | **1.47e-03** | 5.10<br>± 5.37 | 4.39<br>± 4.97 | **0.03** |
| Arousal | -0.18<br>± 0.44 | 0.13<br>± 0.41 | **5.57e-68** | 0.01<br>± 0.45 | 0.13<br>± 0.41 | **5.15e-07** | 0.05<br>± 0.44 | 0.14<br>± 0.44 | **1.42e-03** |
| Valence | -0.003<br>± 0.36 | 0.18<br>± 0.39 | **3.18e-35** | 0.11<br>± 0.41 | 0.17<br>± 0.39 | **3.07e-03** | 0.15<br>± 0.40 | 0.21<br>± 0.40 | **0.01** |

Mean value comparison between case and control groups at acute (up to 4 weeks), ongoing (4-12 weeks after diagnosis), and post-COVID (over 12 weeks after diagnosis) time points. There is no significant difference between case and control groups in any metrics in the period 8 weeks to 4 weeks before diagnosis. Resting heart rate (RHR), sleep efficiency, sleep duration, and step count were calculated relative to a baseline level taken as a mean 12 weeks prior to diagnosis. Uncorrected p-values are reported, but emboldened p-values indicate significance after Benjamini/Hochberg correction.

Considering first the passive metrics, resting heart rate significantly increased in the COVID-19 positive case group compared to the control group in every period. The difference in heart rate in the acute phase (0.58bpm) is less than the following ‘ongoing’ period (1.1bpm) and similar to the post-COVID syndrome period after twelve weeks (0.46bpm). This apparent subdued change during the acute infection is because taking the mean does not properly reflect the non-monotonic changes to resting heart rate during this period. The general group-wide pattern is a peak during the first week of infection, followed by a trough in heart rate during the second week, and finally another, longer lasting, increase (Figure 1). This may imply two acute sub-phases on a shorter timescale than 4 weeks. Step count is also significantly negatively affected during the acute period, but is not significantly changed afterwards. The two sleep metrics show an increase in sleep duration and decrease in efficiency in both case and control groups. There is not a significant difference in duration, but sleep efficiency is significantly decreased throughout all three periods.

**Figure 1:**
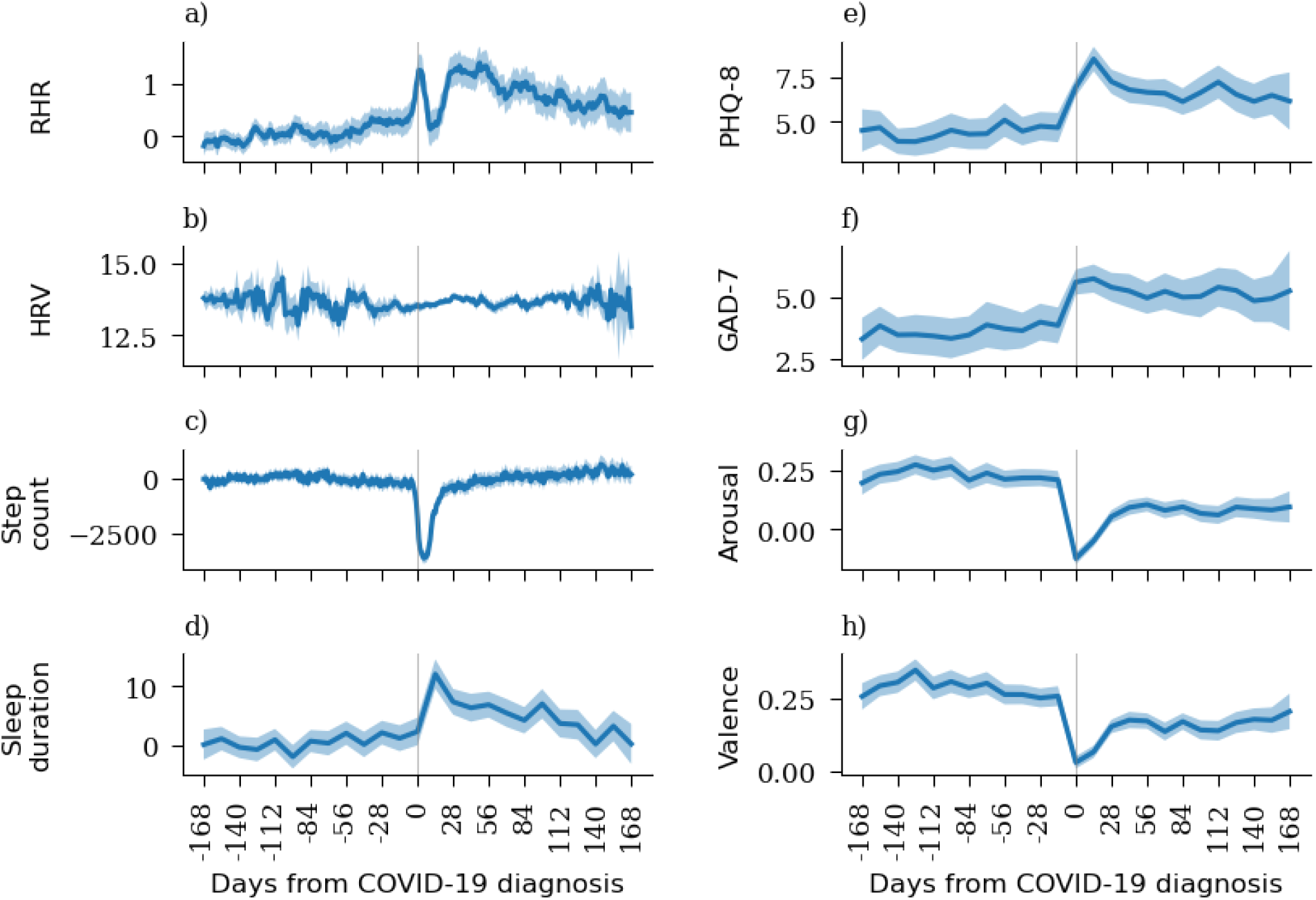
Passive and self-reported measures of mental health across the COVID positive cohort Dates range from 16 weeks prior to 24 weeks post diagnosis of COVID-19. The shaded area corresponds to the 95% confidence interval taken over 14-day windows. a) Daily resting heart rate provided through Fitbit Web API. b) Heart rate variability. c) Change to daily step count from baseline. d) Change in sleep duration from baseline. e) PHQ-8 and f) GAD-7 scores. g) Arousal and h) valence scores, which are reported on a visual analogue scale and range from -1 to +1. HRV: Heart rate variability. RHR: Resting heart rate

All of the self-reported measures of mental health were significantly negatively affected during every period. The mean difference between case and controls for each mental health metric did decrease over time. For example, from a +2.74 (<4 weeks) to +0.98 (>12 week) difference in the average PHQ-8 score. The increased average level and high variance suggests a subset of people suffer persistent symptoms of depression, anxiety, and fatigue (inferred from arousal) for at least twelve weeks.

### Risk factors for LCOVID

#### LCOVID through passive wearable data

A multiple linear regression (Table 4) was carried out to determine whether pre-diagnosis historic fitness wearable data could be used as a risk factor for persistent elevated RHR at 12 weeks post diagnosis (as a proxy for LCOVID). Participants were included if a baseline RHR could be determined in the month prior to diagnosis, if there was at least 4 days of RHR data in the twelfth week, and if they had at least 7 days worth of sleep duration and activity data in the ‘historic’ period. In total, 597 participants had passive data available across the whole time period. The outcome variable was the change in resting heart rate between baseline (−4 to -1 weeks) and 12-weeks post-diagnosis among the COVID-positive cohort. The change in RHR between the baseline and acute phase was included as an independent variable to account for the initial change. That is, given a certain change in the acute phase, what variables are significantly associated with that change persisting. Greater historic activity, the mean duration of time spent taking part in heavy activity between one and two years prior to diagnosis, was negatively correlated with an increase in the outcome variable. These results suggest a slight protective effect against LCOVID for younger and more active people. Female sex was positively associated with persistent LCOVID but not significantly so.

**Table 4:** Risk factor regression of Long-COVID based on passive wearable data

| Variable | Coefficient | Std Err | p-value | [0.025 0.975] CI |
| --- | --- | --- | --- | --- |
| Intercept | -1.9786 | 1.078 | 0.067 | [-4.097 0.139] |
| Age | 0.0274 | 0.011 | **0.017 \*** | [ 0.005 0.050] |
| Female sex | 0.5577 | 0.337 | 0.098 | [-0.104 1.219] |
| Historic activity | -0.0166 | 0.007 | **0.015 \*** | [-0.030 -0.003] |
| Historic sleep | 0.0027 | 0.002 | 0.160 | [-0.001 0.007] |
| Acute ΔRHR | 0.2805 | 0.039 | **1.80e-12\*** | [ 0.204 0.357] |

A linear regression of change in heart rate between baseline and 12 weeks post-diagnosis against age, sex, historic activity, historic sleep, and change in resting heart rate between baseline and acute. P-values are reported uncorrected, but significant results after Benjamini-Hochberg correction are emboldened and marked with an '\*\*'

#### Long COVID through self-reported symptoms

Self-reported symptoms data are visualised using a heatmap in Figure 2. The colourbar on 0.10, the heatmap represents counts (number of reports) while the severity is denoted by the 3 levels (mild, moderate, severe) on the right y-axis. While most symptoms have highest counts and severity around the diagnosis, some symptoms, such as fatigue, persist for longer with a moderate to high severity. Cough and breathing problems also persist for longer periods but with mild severity.

**Figure 2:**
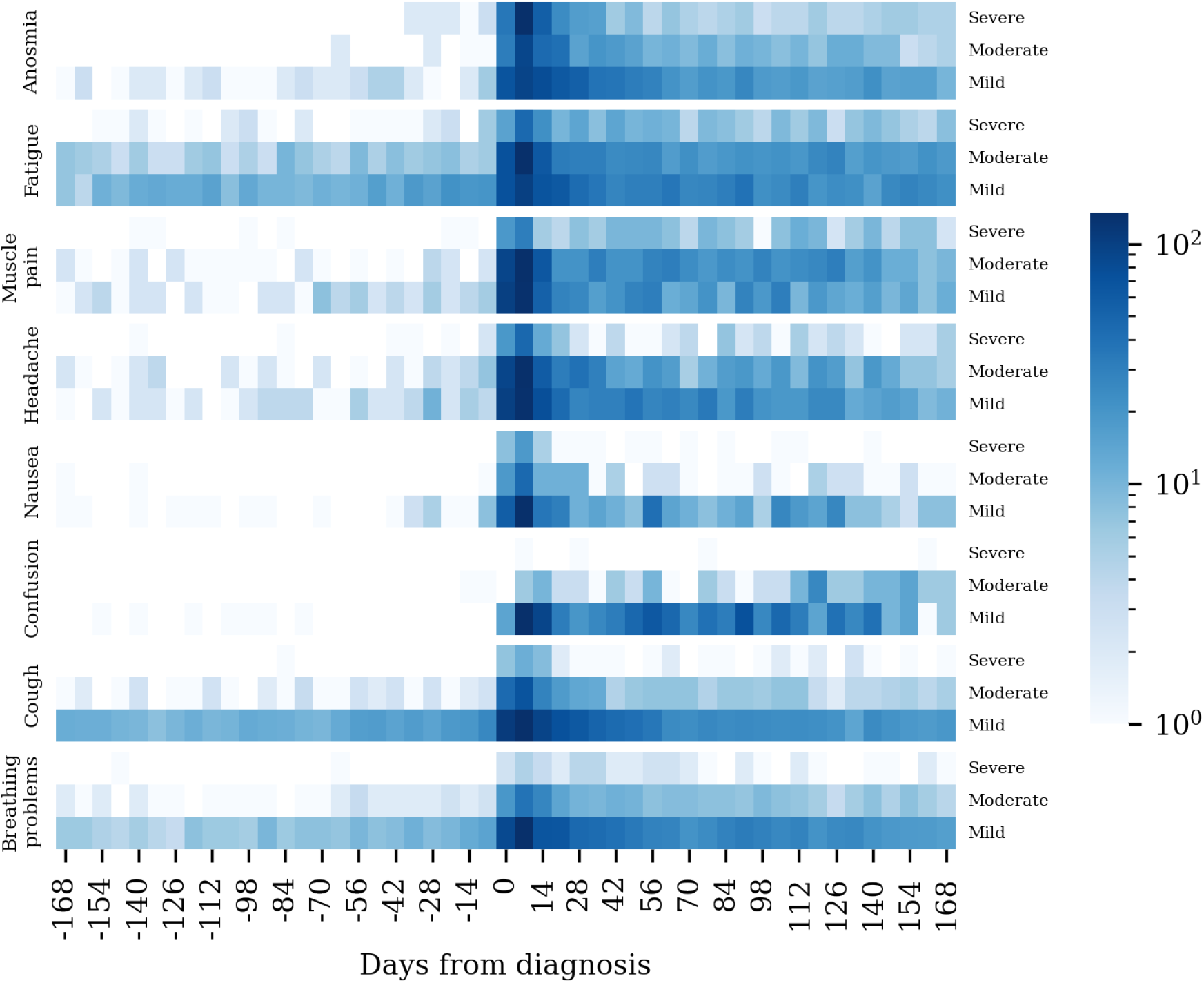
Self-reported symptom heatmap A heatmap showing counts of self-reported symptom severity among the COVID positive cohort around the date of diagnosis. Counts are displayed on a log scale.

To further explore how chronic or acute symptoms of COVID-19 and subsequently how these may relate to LCOVID, durations of various symptoms were plotted (Figure 3). This shows fatigue is typically the longest-lasting symptom, with some exceptional cases reporting fatigue for more than 140 days. The duration of the combined category of any symptom shows that some participants have symptoms lasting over 300 days.

**Figure 3:**
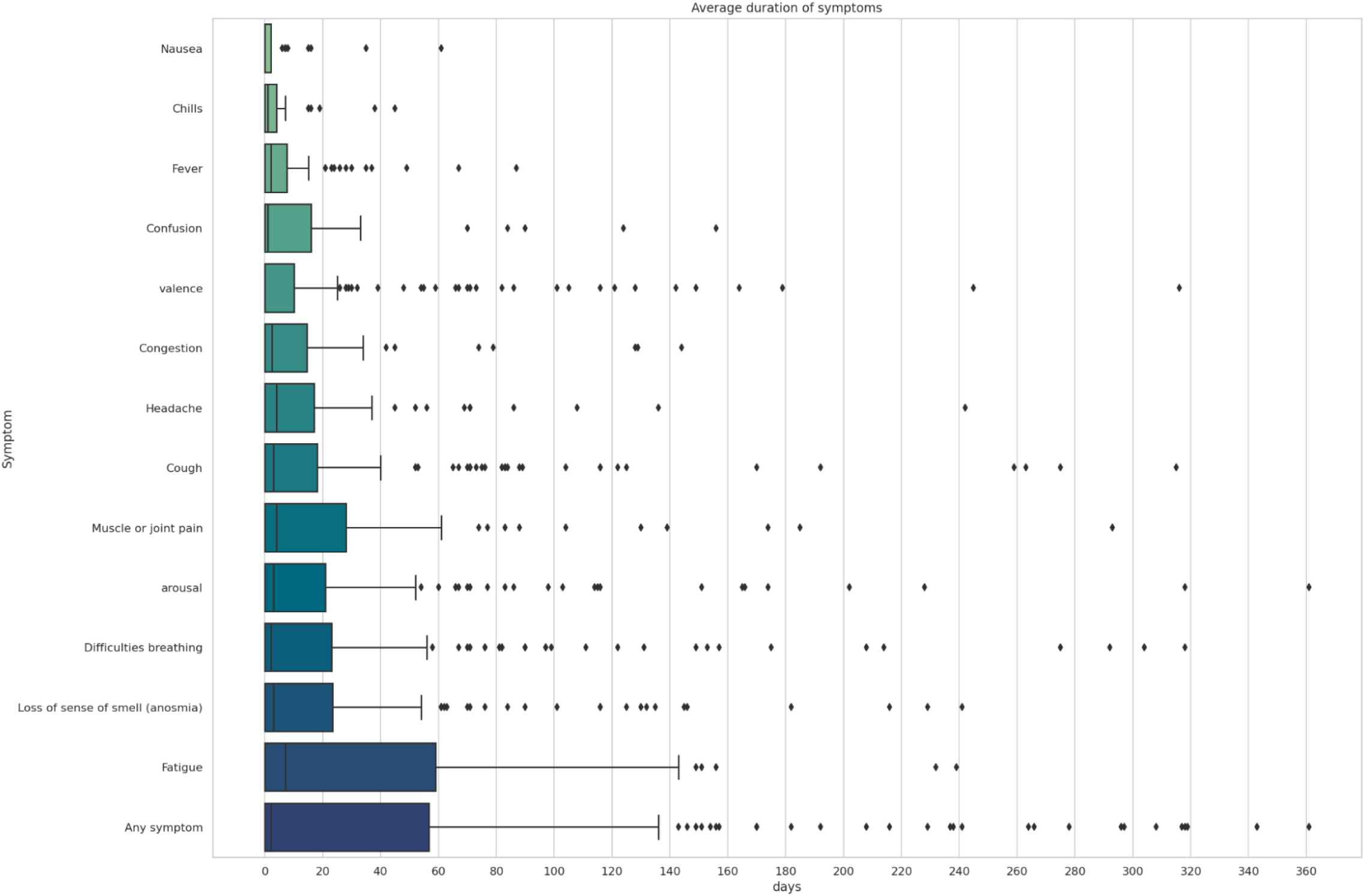
Self-reported symptom duration A box plot representing average duration of symptoms (from the diagnosis date) for participants diagnosed with COVID-19. For clarity 13 most reported symptoms are included. The Any Symptom is an occurrence of any one of the symptoms. LS

### Symptom stratification

As discussed in the methods section, the *L*_*symp*_ and *S*_*symp*_ groups were derived using symptom data, with *L*_*symp*_ participants defined as having reported persistent symptoms for at least 12 weeks. Of the 1327 total diagnosis reported 12.13% (161) reported persistent symptoms and were classified as *L*_*symp*_, with the remaining participants (1104, 83.19%) classified as *S*_*symp*_.

Stratification of the symptom-based cohorts over various socio-demographic factors was carried out (Table 5). There is a significant difference in age between the two cohorts, with the *L*_*symp*_ cohort associated with the older age group. The percentage of participants with certain comorbidities, including asthma, hypertension, diabetes and depression, was higher in the *L*_*symp*_ group, but not significantly so. A higher percentage of people in the *L*_*symp*_ group were employed in part-time roles or were retired while the *S*_*symp*_ cohort had a higher percentage of participants with full-time employment. *L*_*symp*_ also had a higher percentage of participants who were married and had children compared to *S*_*symp*_. No differences can be seen in smoking between the two cohorts.

**Table 5:** Sociodemographic stratification for S-LCOVID and S-SCOVID cohorts

| Variable | Short Covid (>3d) | Long Covid (>12w) |
| --- | --- | --- |
| **General demographics** |  |  |
| Gender [%F, %M] | [76.23, 23.03] | [75.16, 24.84] |
| Age (Mean, std, [IQR]) | 43.85 ± 12.52 [34.0 53.0] | 49.51 ± 10.47 [41.3 57.8] |
| Bmi (Mean, std, [IQR]) | 27.26 ± 4.84 [23.5 30.9] | 28.29 ± 5.48 [24.3 32.9] |
| **Mental Health** |  |  |
| Depression (%) | 19.87 | 25.66 |
| Anxiety, nerves, or generalised anxiety disorder (%) | 8.27 | 6.64 |
| **Physical Health** |  |  |
| Asthma (%) | 17.01 | 23.45 |
| Hypertension or high blood pressure (%) | 6.04 | 8.41 |
| Obesity (%) | 3.66 | 3.98 |
| Diabetes (Type 2) (%) | 2.86 | 4.87 |
| Cancer (%) | 3.66 | 0.44 |
| **Employment** |  |  |
| Full time (%) | 54.21 | 47.79 |
| Part time (%) | 12.08 | 19.03 |
| Retired (%) | 9.54 | 15.49 |
| Self employed (%) | 5.88 | 7.08 |
| At home carer (%) | 2.54 | 3.54 |
| Unemployed (%) | 2.23 | 2.21 |
| **Marriage and children** |  |  |
| Married (%) | 53.42 | 61.06 |
| Single (%) | 13.67 | 17.7 |
| Living with partner (%) | 13.99 | 10.62 |
| Living apart from partner (%) | 3.34 | 3.1 |
| Divorced (%) | 3.82 | 2.21 |
| Separated (%) | 3.18 | 1.77 |
| Children - y (%) | 65.59 | 74.34 |
| Children - n (%) | 28.46 | 22.12 |
| **Smoking** |  |  |
| Smoker - never (%) | 55.03 | 56.52 |
| Smoker - ex (%) | 30.04 | 31.06 |
| Smoker - 1-10 (%) | 2.93 | 2.48 |
| Smoker - ecig (%) | 3.02 | 2.48 |
| Smoker - 11-20 (%) | 1.46 | 1.86 |
| Smoker - <1 (%) | 1.74 | 0.62 |

To visualise the differences between the *S*_*symp*_ and *L*_*symp*_ cohorts around diagnosis, various metrics were plotted as shown in Figure 4. Resting Heart Rate (RHR) was higher in the *L*_*symp*_ cohort and stayed high for a longer period. The pattern of elevated and reduced heart rate in the acute phase are also more pronounced in the *L*_*symp*_ cohort. There appear to be some inherent differences in step count between *L*_*symp*_ and *S*_*symp*_ (<100 days before). Interestingly, the *L*_*symp*_ cohort has a greater step count and the difference in the rate of drop in steps approaching the diagnosis date, whereas the recovery of steps appear more similar. Sleep duration was higher in the *S*_*symp*_ cohorts over the whole time period but the difference in durations between *L*_*symp*_ and *S*_*symp*_ peaked close to the diagnosis date.

**Figure 4:**
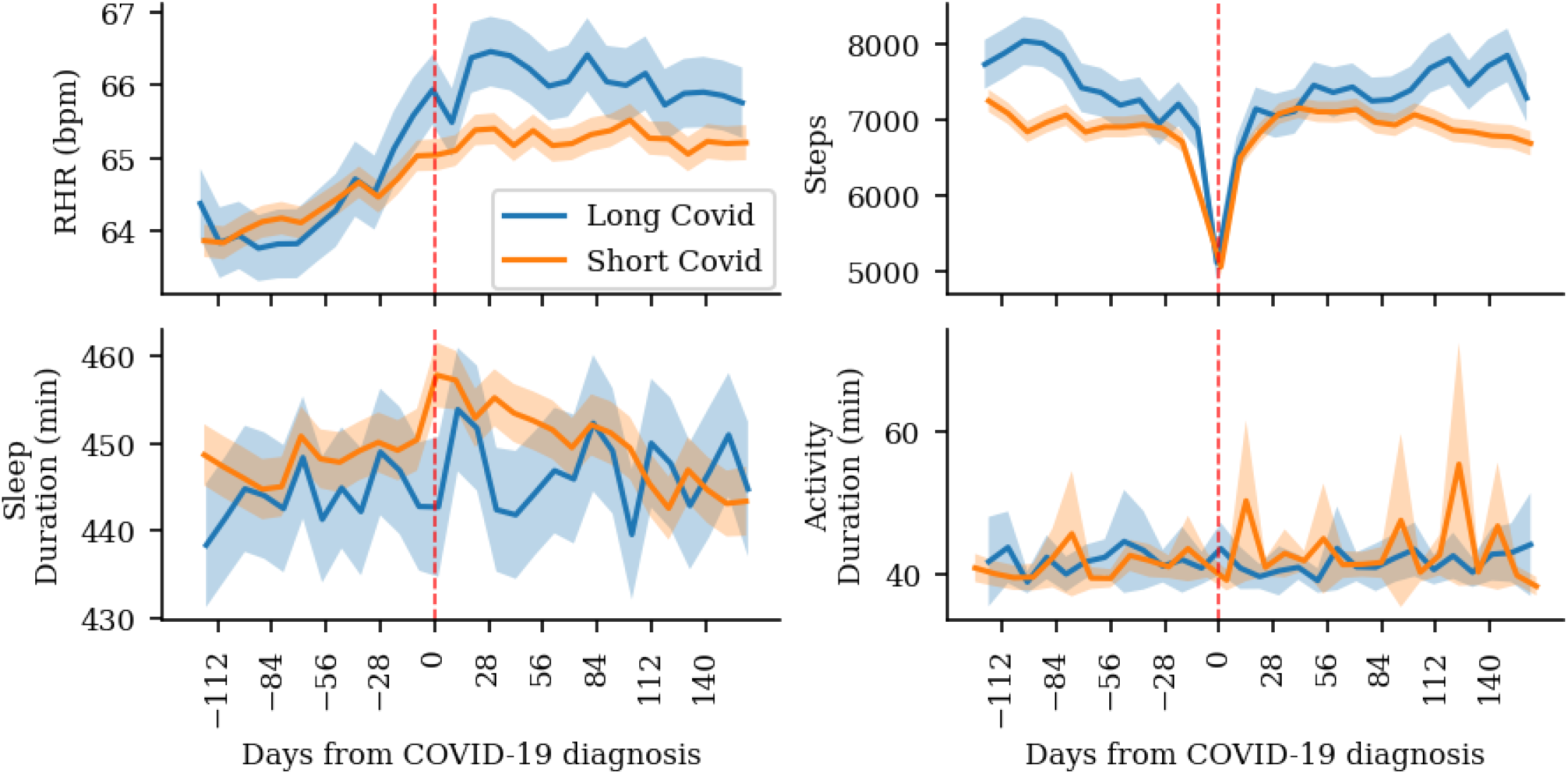
Passive metrics across symptom-based short and LCOVID cohorts Comparison of passive and self-reported symptom measures for *S*_*symp*_ and *L*_*symp*_ cohorts, ranging from 16 weeks prior to 24 weeks post diagnosis of COVID-19. The shaded area corresponds to the 95% confidence interval taken over 10-day windows.

A series of Multiple Logistic Regressions were performed to establish the likelihood of having LCOVID based on the effects of sociodemographic, wearable, and mental health survey covariates. Each regression was on a single explanatory variable and adjusted for age, gender and ethnicity. The p-values were adjusted using the Benjamini-Hochberg correction for multiple testing.

Features for various independent variables were calculated based on the mean value over the year prior to diagnosis, the mean value during the acute phase (diagnosis + 14 days). Of the 1327 participants with a positive COVID diagnosis, 1213 [1060 *S*_*symp*_ and 153 *L*_*symp*_] were included after exclusion of participants with missing data for the required variables. The inclusion criteria for continuous variables was based on a completion rate of at least 60% in the baseline and acute phases. More participants may have been excluded in different regressions because of missing data per variable.

The logistic regression results are given in Table 6. The significant Benjamini-Hochberg corrected p-values (p-value < 0.05) are marked with a * and in bold. Furthermore, a forest plot was generated to visualise the effects of the variables (Figure 5) with significant effects shown in orange. Age ranges were used to assess the effect of age on developing LCOVID and these were the most prominent risk factors for LCOVID. Age Range (10-30) was used as the reference for other categories. All other age ranges had a significant effect with a rise in the odds ratio at each level, suggesting that the odds of developing LCOVID increases with an increase in age. The 50-60 age group was 6.5 times more likely and the oldest group (60+) was 6.4 more likely to have LCOVID than the reference group. Female gender (with reference as male) did not show a significant effect. Comorbidities, such as asthma, hypertension, and diabetes, did not show a significant effect (p-value < 0.05) on the presence of LCOVID, but the odds ratio were greater than one. Passive features and self-reported questionnaires were also considered as risk factors for LCOVID. Acute and Baseline phase PHQ8 scores had a significant effect.

**Table 6:** Multiple logistic regression for *L*_*symp*_.

| Dependent Variable | OR | P-value * | 2.5% | 97.5% |
| --- | --- | --- | --- | --- |
| Age | 1.03037 | **0.01113\*** | 1.01262 | 1.04844 |
| Bmi | 1.01509 | 0.47034 | 0.98541 | 1.04567 |
| Asthma | 1.35638 | 0.37109 | 0.83104 | 2.21382 |
| Hypertension | 1.68335 | 0.29906 | 0.82024 | 3.45471 |
| Diabetes | 1.35638 | 0.37109 | 0.83104 | 2.21382 |
| Gender Female (reference = Male) | 1.08099 | 0.8061 | 0.66169 | 1.766 |
| Age Range [30-40] (reference = [10-30]) | 3.4188 | 0.16243 | 0.9475 | 12.33587 |
| Age Range [40-50] (reference = [10-30]) | 5.43478 | **0.04354\*** | 1.57941 | 18.70117 |
| Age Range [50-60] (reference = [10-30]) | 6.57051 | **0.02487\*** | 1.94017 | 22.25143 |
| Age Range Above 60 (reference = [10-30]) | 6.41026 | **0.02745\*** | 1.83102 | 22.44179 |
| Ethnicity Others (black, asian, others) (reference=White) | 0.97608 | 0.96288 | 0.35207 | 2.70609 |
| Current Smoker (reference = never) | 0.85398 | 0.8061 | 0.35404 | 2.05987 |
| Ex Smoker (reference = never) | 0.89694 | 0.8061 | 0.56322 | 1.42839 |
| Baseline steps | 0.99999 | 0.8061 | 0.99993 | 1.00005 |
| Acute phase steps | 0.99997 | 0.47034 | 0.99991 | 1.00003 |
| Baseline resting heart rate | 1.03403 | 0.20396 | 0.99418 | 1.07547 |
| Acute phase resting heart rate | 1.03569 | 0.07045 | 1.00583 | 1.06644 |
| Baseline activity minutes | 0.99497 | 0.5177 | 0.98343 | 1.00665 |
| Acute phase activity minutes | 1.001 | 0.8061 | 0.99404 | 1.00801 |
| Baseline sleep minutes | 1.00386 | 0.15988 | 1.00003 | 1.00771 |
| Acute phase sleep minutes | 1.00165 | 0.47814 | 0.99818 | 1.00514 |
| Baseline sleep efficiency | 0.99664 | 0.8061 | 0.97753 | 1.01612 |
| Acute phase sleep efficiency | 0.98888 | 0.47034 | 0.96697 | 1.01129 |
| Baseline PHQ8 score | 1.09338 | **0.04436\*** | 1.02267 | 1.16898 |
| Acute phase PHQ8 score | 1.09489 | **0.00301\*** | 1.046 | 1.14606 |
| Baseline GAD7 score | 1.05617 | 0.29906 | 0.97874 | 1.13972 |
| Acute phase GAD7 score | 1.06283 | 0.05935 | 1.01249 | 1.11568 |
| Baseline symptom severity | 1.26064 | 0.16243 | 0.98573 | 1.61222 |
| Acute phase symptom severity | 0.93129 | 0.20396 | 0.85717 | 1.01181 |
| Acute phase number of symptoms | 0.86327 | 0.16243 | 0.74269 | 1.00342 |

The values in bold and marked with \* show that the variable has a significant effect on the outcome of having long COVID.

**Figure 5:**
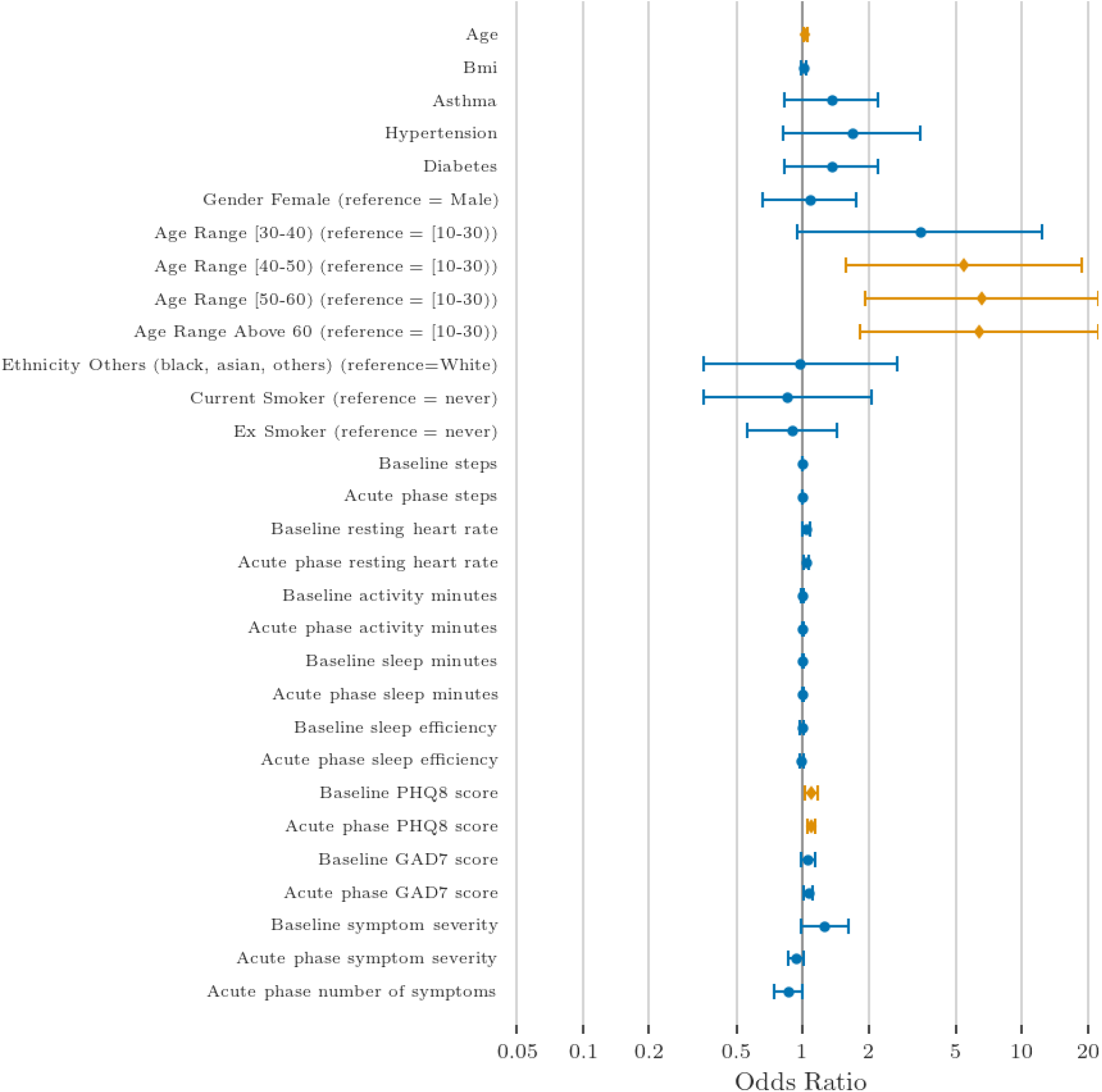
Logistic regression odds ratio for variables across symptom-based short and long COVID cohorts Odds Ratio with 95% confidence interval, adjusted for age. The red colours show that the dependent variable has a significant effect (p-value < 0.05) on the independent variable.

## Discussion

In this study we investigate persistent symptoms of and recovery from COVID-19 through the lens of mobile health data. We find a signal for LCOVID in both passive and active metrics, as well as associates to comorbidities, physiological metrics, and prior behaviour.

At a group-wide level, several wearable and mental health metrics are significantly changed during the acute COVID infection, some of which remain significantly different from the control group for longer than twelve weeks (Figure 1). Resting heart rate is the wearable metric with the longest lasting noticeable change. We estimate the proportion of participants with a long-term change to their heart rate coinciding with COVID-19 infection to be 7% on the basis of the BSTS model fit of heart rate data prior to infection, somewhat less than another study in which 13.4% of participants had a RHR of five bpm or more at twelve weeks [20]. Depression, anxiety, and self-rated arousal-valence remain negatively affected in the LCOVID phase.

As has previously been reported[20, 21], we observe a pattern of transient RHR elevation from COVID-19 infection onset for a week, followed by a period of transient reduced RHR from the second to third week, and finally a chronic or long-lasting increase in resting heart rate in some participants which can last several months or longer (Figure 4. This pattern of acute elevation and subsequent reduction is more prominent in the LCOVID cohort compared to the short COVID cohort, as shown in Figure 4.

The changing nature of the pandemic not only led to a changing understanding of what should be monitored in a study of this type, but importantly led to large societal and public health interventions[22]. Many of those will also have had an effect on mental well-being and physical health. For instance, lockdown measures coincide with increased infection levels and also have an effect on activity[23, 24], sleep[25, 26], heart rate[26], and mental health[25, 27]. Therefore, when monitoring recovery in COVID-19 through mobile health, we should also consider the wider societal context. By time matching the control group we try to account for those changes, however, the impact of events concurrent to COVID-19 infection could be investigated in more detail.

An advantage of requesting existing wearable data from users of commercial fitness wearables is the ability to create a longitudinal dataset covering a period prior to enrolment, in some cases for many years. A higher level of historic physical activity is negatively correlated with development of the passive marker of LCOVID, the persistent increased RHR at twelve weeks post-diagnosis. To our knowledge, no other study considers the effect of historic activity or fitness level on LCOVID, but it has been demonstrated to reduce the severity and risk of hospitalisation in acute COVID[28]. Sleep duration was not significantly associated, but may be worth further investigation alongside other markers of historic health in larger datasets. Age was significantly positively associated, in agreement with multiple other studies[3, 29, 30] and the symptom-based findings in this study. Female sex was not significantly different to male sex, but did have a positive coefficient and a fairly low p-value. It seems likely that in a larger or more powerful dataset, female sex would be a risk factor for LCOVID, in line with previously published research[3, 29–31].

### Symptom Findings

The estimated prevalence of LCOVID in the literature is diverse. Our finding of the proportion of people in the *L*_*symp*_ group, after reporting a diagnosis and having persistent symptoms for 12 weeks (12.13%), is consistent with results from UK government Office for National Statistics (13.7%) in a study involving 20,000 participants[32]. However, studies have reported different prevalence rates for LCOVID at twelve weeks, from 2.6%[30] to 14.8% [33] and 37% [3]. Variance could be explained through sociodemographic differences across cohorts, methodological differences in the collection of symptom data, or how LCOVID is defined based on collected symptom data.

Our results show fatigue is the longest lasting symptom, with several participants experiencing fatigue for more than 140 days, which is consistent with previously published research[3, 30, 34, 35].

In agreement with previous studies[3, 29, 30] and the RHR-based regression, age was found to be a significant risk factor for LCOVID (*L*_*symp*_ cohort) with ages greater than 50 at very high risk. BMI (and obesity) was not a significant factor in our study. Other studies have shown that the female sex had a positive association with LCOVID[3, 29–31] but this was inconclusive in our study. A lack of power in this cohort reduces our ability to educe significance. Comorbidities like asthma, hypertension, and diabetes are potential risk factors for LCOVID, with non-significant p-values, which would agree with previous findings[29, 30]. This is not conclusive, as the *L*_*symp*_ cohort was defined through persistent symptoms which could also be caused by chronic illnesses, not necessarily COVID-19.

Investigation of passive metrics from wearables and self-reported questionnaires revealed that while a depression comorbidity was not significantly associated, the average PHQ8 score over the year prior to a COVID-19 diagnosis and during the acute phase of the disease was positively associated with LCOVID, indicating that a period of low mood before and during the disease could be a risk factor for LCOVID. Further resting heart rate in the acute phase of the disease also had a positive relation to LCOVID, with a low but non-significant p-value of 0.07, and could be a potential risk factor. The persistently increased RHR in the *L*_*symp*_ cohort, as visualised in Figure 4, also demonstrates a level of coherence between the two approaches to determining LCOVID.

### Strengths and weaknesses

This study brings together COVID-19 self-reported symptoms, passive wearable data, and regular mental health surveys in a population who are not necessarily hospitalised. Both the symptom and passive approaches demonstrate that age is a risk factor for LCOVID. The availability of historic wearable data enabled the development of a long duration baseline with which to compare subsequent changes during COVID-19 infection. Increased historic activity, which suggests a participant who had previously engaged in more exercise, is protective against the passive-based proxy for LCOVID.

The passive data approach uses existing data which is highly available among those who own wearable devices, is unbiased by subjective rating and identification of symptoms, and doesn’t burden participants, but is limited in symptom scope to what a wearable device can measure. Meanwhile, the symptom self-report based methodology allows reporting of a wider range of symptoms than would be captured through wearable sensors and more concrete labels in the absence of a robust LCOVID classification algorithm for passive data, but relies on engaged and persistent participants.

There are multiple limitations to this study. Firstly, the definition of a LCOVID group on the basis of self-reported symptoms or by using resting heart rate as a proxy measure are weak approximations of a true diagnosis label. While we were able to show group-wide differences during the post-COVID period in self-reported mental health measures and passive wearable device biosignals, the use of the change in resting heart rate in the post-COVID period is non-specific and the effect of COVID-19 can be overwhelmed by natural variability in the individual case. Self-reported symptom monitoring requires time and commitment on the part of the person monitoring their COVID-19 recovery, which may be unrealistic to expect, especially given the increased prevalence of depression and fatigue. In both cases we assume a consistent deviation from a healthy baseline. However, symptoms of LCOVID may fluctuate or show signs of relapse and remission. Developing a model to identify LCOVID in mobile health data using another, labelled, dataset to then stratify COVID-19 recovery in datasets without explicit labelling may be an approach worth following, with a similar approach demonstrated recently in an electronic health records study of LCOVID[36].

Secondly, the nature of mobile health studies in general and of a community-sourced study, which relies on motivation and interest by participants, can lead to sporadic completion rates. Data completeness is reliant on what a participant is able and willing to share and on their continued engagement with the study. Meanwhile, the open remote enrolment paradigm biases the groups participating to those who have the studies published in a way that reaches them and that are self-motivated to take part. For example, the proportion of female participants is notably higher, a pattern that is seen across similar studies [24, 37]. This may be partially addressed through larger-scale studies or a meta-analysis including the similar studies that are running across various countries.

## Conclusion

In conclusion, we demonstrate a measurable difference in measures of mental wellbeing and in biosignals from commercial wearable devices between COVID-19 positive and non-diagnosed participants during the sixteen weeks following diagnosis. Two methods of inferring the presence of LCOVID are compared. One method is based on persistent changes in resting heart rate and the other on persistent self-reported symptoms of COVID-19. For the self-reported symptoms method, the LCOVID risk factors were explored using demographics, self-reports and passive wearable data, and compared with results from existing literature. In the future we plan to assess the feasibility of combining studies to create larger datasets or meta-analyses, to develop a LCOVID detection algorithm for use in mobile health data, and to investigate the additional effect of public health and safety measures.

## Data Availability

Data from participants who consented to share an anonymised copy are available on reasonable request to the authors.

## Notes

### Competing Interest Statement

The authors have declared no competing interest.

### Funding Statement

This study was supported by the NIHR Maudsley
Biomedical Research Centre at South London and Maudsley NHS Foundation Trust and Kings College London but did not receive specific funding.

### Author Declarations

PNM Research Ethics Panel of King's College London gave ethical approval for this work

## References

1. H. Ritchie et al.: Coronavirus Pandemic (COVID-19). Our World in Data (2020). https://ourworldindata.org/coronavirus.

2. P. H. Roth and M. Gadebusch-Bondio: The contested meaning of long COVID Patients, doctors, and the politics of subjective evidence. Social Science &: Medicine 292 (Jan. 2022), 114619. doi: 10.1016/j.socscimed.2021.114619.

3. M. Whitaker et al.: Persistent COVID-19 symptoms in a community study of 606,434 people in England. Nature Communications 13(1) (Apr. 2022). doi: 10.1038/s41467-022-29521-z.

4. D. Munblit et al.: Long COVID: aiming for a consensus. The Lancet Respiratory Medicine 10(7) (July 2022), 632–634. doi: 10.1016/s2213-2600(22)00135-7.

5. COVID-19 rapid guideline: managing the long-term effects of COVID-19. Mar. 2022. url: https://www.nice.org.uk/guidance/ng188/resources/covid19-rapid-guideline-managing-the-longterm-effects-of-covid19-pdf-51035515742 (visited on 09/19/2022).

6. T. M. Schou, S. Joca, G. Wegener, and C. Bay-Richter: Psychiatric and neuropsychiatric sequelae of COVID-19 A systematic review. Brain, Behavior, and Immunity 97 (Oct. 2021), 328–348. doi: 10.1016/j.bbi.2021.07.018.

7. M. Gavriatopoulou et al.: Organ-specific manifestations of COVID-19 infection. Clinical and Experimental Medicine 20(4) (July 2020), 493–506. doi: 10.1007/s10238-020-00648-x.

8. A. Mezlini et al.: Estimating the Burden of Influenza-like Illness on Daily Activity at the Population Scale Using Commercial Wearable Sensors. JAMA Network Open 5(5) (May 2022), e2211958. doi: 10.1001/jamanetworkopen.2022.11958.

9. C. Stewart et al.: Investigating the Use of Digital Health Technology to Monitor COVID-19 and Its Effects: Protocol for an Observational Study (Covid Collab Study). JMIR Research Protocols 10(12) (Dec. 2021), e32587. doi: 10.2196/32587.

10. K. Kroenke et al.: The PHQ-8 as a measure of current depression in the general population. Journal of Affective Disorders 114(1-3) (Apr. 2009), 163–173. doi: 10.1016/j.jad.2008.06.026.

11. R. L. Spitzer, K. Kroenke, J. B. W. Williams, and B. Löwe: A Brief Measure for Assessing Generalized Anxiety Disorder. Archives of Internal Medicine 166(10) (May 2006), 1092. doi: 10.1001/archinte.166.10.1092.

12. J. A. Russell: A circumplex model of affect. Journal of Personality and Social Psychology 39(6) (Dec. 1980), 1161–1178. doi: 10.1037/h0077714.

13. A. Russell, C. Heneghan, and S. Venkatraman: Investigation of an estimate of daily resting heart rate using a consumer wearable device (Oct. 2019). doi: 10.1101/19008771.

14. A. Natarajan, A. Pantelopoulos, H. Emir-Farinas, and P. Natarajan: Heart rate variability with photoplethysmography in 8 million individuals: a cross-sectional study. The Lancet Digital Health 2(12) (Dec. 2020), e650–e657. doi: 10.1016/s2589-7500(20)30246-6.

15. Web API. 2022. url: https://dev.fitbit.com/build/reference/web-api/ (visited on 09/19/2022).

16. S. Seabold and J. Perktold: Statsmodels: Econometric and Statistical Modeling with Python. Proceedings of the Python in Science Conference. SciPy, 2010. doi: 10.25080/majora-92bf1922-011.

17. K. H. Brodersen, F. Gallusser, J. Koehler, N. Remy, and S. L. Scott: Inferring causal impact using Bayesian structural time-series models. The Annals of Applied Statistics 9(1) (Mar. 2015). doi: 10.1214/14-aoas788.

18. E. Brunner and U. Munzel: The Nonparametric Behrens-Fisher Problem: Asymptotic Theory and a Small-Sample Approximation. Biometrical Journal 42(1) (2000), 17–25. doi: https://doi.org/10.1002/(SICI)1521-4036(200001)42:1<17::AID-BIMJ17>3.0.CO;2-U.

19. Y. Benjamini and Y. Hochberg: Controlling the False Discovery Rate: A Practical and Powerful Approach to Multiple Testing. Journal of the Royal Statistical Society: Series B (Methodological) 57(1) (Jan. 1995), 289–300. doi: 10.1111/j.2517-6161.1995.tb02031.x.

20. J. M. Radin et al.: Assessment of Prolonged Physiological and Behavioral Changes Associated With COVID-19 Infection. JAMA Network Open 4(7) (July 2021), e2115959. doi: 10.1001/jamanetworkopen.2021.15959.

21. A. Natarajan, H.-W. Su, and C. Heneghan: Occurrence of Relative Bradycardia and Relative Tachycardia in Individuals Diagnosed With COVID-19. Frontiers in Physiology 13 (May 2022). doi: 10.3389/fphys.2022.898251.

22. B. T. Snoeijer, M. Burger, S. Sun, R. J. B. Dobson, and A. A. Folarin: Measuring the effect of Non-Pharmaceutical Interventions (NPIs) on mobility during the COVID-19 pandemic using global mobility data. npj Digital Medicine 4(1) (May 2021). doi: 10.1038/s41746-021-00451-2.

23. B. Constandt et al.: Exercising in Times of Lockdown: An Analysis of the Impact of COVID-19 on Levels and Patterns of Exercise among Adults in Belgium. International Journal of Environmental Research and Public Health 17(11) (June 2020), 4144. doi: 10.3390/ijerph17114144.

24. S. Sun et al.: Using Smartphones and Wearable Devices to Monitor Behavioral Changes During COVID-19. Journal of Medical Internet Research 22(9) (Sept. 2020), e19992. doi: 10.2196/19992.

25. A. S. Kochhar et al.: Lockdown of 1.3 billion people in India during Covid-19 pandemic: A survey of its impact on mental health. Asian Journal of Psychiatry 54 (Dec. 2020), 102213. doi: 10.1016/j.ajp.2020.102213.

26. J. L. Ong, T. Lau, M. Karsikas, H. Kinnunen, and M. W. L. Chee: A longitudinal analysis of COVID-19 lockdown stringency on sleep and resting heart rate measures across 20 countries. Scientific Reports 11(1) (July 2021). doi: 10.1038/s41598-021-93924-z.

27. K. F. Ahrens et al.: Differential impact of COVID-related lockdown on mental health in Germany. World Psychiatry 20(1) (Jan. 2021), 140–141. doi: 10.1002/wps.20830.

28. J. P. Brandenburg, I. A. Lesser, C. J. Thomson, and L. V. Giles: Does Higher Self-Reported Cardiorespiratory Fitness Reduce the Odds of Hospitalization From COVID-19? Journal of Physical Activity and Health 18(7) (July 2021), 782–788. doi: 10.1123/jpah.2020-0817.

29. A. Subramanian et al.: Symptoms and risk factors for long COVID in non-hospitalized adults. Nature Medicine 28(8) (July 2022), 1706–1714. doi: 10.1038/s41591-022-01909-w.

30. C. H. Sudre et al.: Attributes and predictors of long COVID. Nature Medicine 27(4) (Mar. 2021), 626–631. doi: 10.1038/s41591-021-01292-y.

31. R. A. Evans et al.: Clinical characteristics with inflammation profiling of long COVID and association with 1-year recovery following hospitalisation in the UK: a prospective observational study. The Lancet Respiratory Medicine 10(8) (Aug. 2022), 761–775. doi: 10.1016/s2213-2600(22)00127-8.

32. Prevalence of ongoing symptoms following coronavirus (COVID-19) infection in the UK: 1 April 2021. Apr. 2021. url: https://www.ons.gov.uk/peoplepopulationandcommunity/healthandsocialcare/conditionsanddiseases/bulletins/prevalenceofongoingsymptomsfollowingcoronaviruscovid19infectionintheuk/1april2021 (visited on 09/19/2022).

33. E. T. Cirulli et al.: Long-term COVID-19 symptoms in a large unselected population (Oct. 2020). doi: 10.1101/2020.10.07.20208702.

34. H. E. Davis et al.: Characterizing long COVID in an international cohort: 7 months of symptoms and their impact. eClinicalMedicine 38 (Aug. 2021), 101019. doi: 10.1016/j.eclinm.2021.101019.

35. Prevalence of ongoing symptoms following coronavirus (COVID-19) infection in the UK: 4 August 2022. Aug. 2022. url: https://www.ons.gov.uk/peoplepopulationandcommunity/healthandsocialcare/conditionsanddiseases/bulletins/prevalenceofongoingsymptomsfollowingcoronaviruscovid19infectionintheuk/4august2022 (visited on 09/19/2022).

36. E. R. Pfaff et al.: Identifying who has long COVID in the USA: a machine learning approach using N3C data. The Lancet Digital Health 4(7) (July 2022), e532–e541. doi: 10.1016/s2589-7500(22)00048-6.

37. S. Reade et al.: Cloudy with a Chance of Pain: Engagement and Subsequent Attrition of Daily Data Entry in a Smartphone Pilot Study Tracking Weather, Disease Severity, and Physical Activity in Patients With Rheumatoid Arthritis. JMIR mHealth and uHealth 5(3) (Mar. 2017), e37. doi: 10.2196/mhealth.6496.

